# Impact of depression on personal hygiene practices- A cross-sectional study among university students in Bangladesh

**DOI:** 10.1101/2025.04.11.25325655

**Authors:** Fouzia Akter, Akibul Islam Chowdhury, Md. Nawal Sarwer

## Abstract

**Background:** This study explores the relationship between depression and personal hygiene practices among university students in Bangladesh.

**Methods:** A cross-sectional online survey was conducted, utilizing an 18-item Personal Hygiene Practice Questionnaire (PHPQ) and the Center for Epidemiologic Studies Depression Scale (CES-D) to assess hygiene behaviors and depression risk among 1,913 undergraduate students in Dhaka. Data were analyzed using chi-square tests and ordered logistic regression. The PHPQ was validated through item analysis, internal consistency, construct validity and reliability tests.

**Results:** A high prevalence of depression risk was revealed with 79.9% of females and 73.9% of males. Females demonstrated superior hygiene practices, with 90.1% classified as having good hygiene compared to 75.0% of males. Accommodation type significantly influenced both depression and hygiene, as students living in privately managed housing exhibited better hygiene practices (88.6% good hygiene) and lower depression risk (73.2%) compared to those living at home (79.2%) or in university housing (78.7%). Ordered logistic regression analysis indicated that students at risk of depression had 65% lower odds of maintaining better hygiene practices (OR = 0.36, p < 0.001), and male students were 68% less likely to have higher hygiene scores than females (OR = 0.32, p < 0.001). The Exploratory Factor Analysis and Cronbach’s alpha are confirming its reliability (α = 0.83) and strong internal consistency.

**Conclusion:** These findings underscore the need for targeted interventions in university settings to address mental health and hygiene education. Further research should explore socio-economic and cultural factors influencing these relationships.

## Introduction

Depression is a common mental health disease that has a substantial influence on people’s daily life, including their personal hygiene habits. As a common yet often under-recognized condition, depression affects millions worldwide, with a notable prevalence among university students (1). This demographic is particularly vulnerable due to the unique stressors associated with academic life, including academic performance, studying in the English language, heavy lecture schedule, pressure to succeed, future planning, and social challenges (2). Other factors associated with mental illness are demographical including gender, residence, relationship status, socioeconomic status, loneliness, personal autonomy, family and peer pressure (2–4).

Personal hygiene is crucial for maintaining overall health, preventing infectious diseases, and promoting psychological and social well-being (5). Individuals experiencing depression frequently struggle with daily self-care routines due to symptoms like low motivation, fatigue, and cognitive difficulties. Neglecting hygiene can further contribute to poor physical health, social isolation, and decreased self-esteem, potentially exacerbating depressive symptoms in a detrimental cycle of declining mental and physical well-being (6,7). The relationship between personal hygiene and depression needs a comprehensive understanding as it is not well established.

At University level, students have experienced a transition to adulthood. They have moved away from their family into new places and cope with new environment (4) which might lead to both positive and negative changes in their lifestyles (8). Some students may adopt a healthier lifestyle while others may struggle with their new environment and academic life which may have an impact on their personal health related activities.

Previous studies highlighted that depression among university students in Bangladesh is a significant concern, with prevalence rates ranging from 28.7% to 47.3% (9). Several factors contribute to depression, including years of study, stressful life events, suicidal attempts, inadequate monthly allowance, substance use, physical and psychological illness, and excessive social media use (10). The prevalence of depression among university students is rising, yet studies exploring its impact on personal hygiene remain limited.

The relationship between personal hygiene and depression is multifaceted. Previous studies at university settings have shown association between depression and unhealthy lifestyles along with disruption in daily routine activities (8,11). Depression can lead to a lack of motivation and energy, making routine care activities feel overwhelming. Some studies showed hygiene related practices at school settings may influence absenteeism and poor academic performance (12) which may also lead to depression. Moreover, poor personal hygiene may have an impact on poor individual health and social responsibilities. Students with poor personal hygiene may face social stigmatization leading to isolation and depression. Despite these concerns, limited empirical research has examined the relationship between depression and hygiene behaviors among university students, highlighting a significant gap in literature. Therefore, addressing personal hygiene within the context of mental health is vital.

To address this gap, our study explores the association between depression and personal hygiene practices among university students in Bangladesh using a novel 18-item Personal Hygiene Practice Questionnaire (PHPQ).

## Methodology

### Study design and Location

We conducted this cross-sectional study among university students using an online survey from Dhaka city due to its significant concentration of universities. Dhaka has the highest number of universities in Bangladesh with a university present in every 5.38 square kilometers. Due to this Dhaka is known as a central hub for higher education in the country (13).

### Sampling and Data Collection

A structured questionnaire was distributed via a Google Form link to ensure accessibility for participants across various locations from September to December 2024. This online method enhances self-disclosure on sensitive topics (Krantz and Reips, 2017). The link was shared across university networks, student groups, and social media to ensure sample diversity. Initially, 2,030 responses were collected, but after applying exclusion criteria for missing values and inconsistencies, the final analytical sample comprised 1,913 valid responses.

### Study Measures

The socio-demographic section contained data on age, gender, type of university, individual’s subject major and their level of study, place of residence, parental education, and family income in Bangladeshi currency (BDT). We categorized family income into four groups using the quartile method.

### The Center for Epidemiologic Studies Depression Scale (CES-D)

In this study, we used CES-D scale to assess depression among university students in Dhaka, Bangladesh, as it has been validated in various population including young adults and in low middle income countries. It is comprised of 20 self-reported items that measure different dimensions of depression. Respondents rate the frequency of each symptom over the past week on a four-point Likert scale, ranging from 0 (rarely or none of the time, less than 1 day) to 3 (most or all the time, 5-7 days). The total score range is 0–60. A score of 16 anc d above was used to define a case of likely depression or at risk of depression and a score less than 16 was defined as not at risk of depression (14).

### Personal Hygiene Practice Questionnaire

We developed a new questionnaire to assess personal hygiene practices among university students, guided by discussions with faculty members from the Departments of Nutrition and Food Engineering, Pharmacy, and Public Health. Expert review led to the refinement of an initial 21-item questionnaire, resulting in an 18-item final version that encompasses various dimensions of personal hygiene, including hand hygiene and personal cleanliness. After developing the questionnaire, a pilot survey was conducted with 44 students to check whether they could easily understand the questions and response options. 93.18% of students didn’t find any difficulties in understanding the question and 86.36% of students had no issues with the options of the questions. We also tested the internal reliability of the questionnaire using Cronbach’s alpha and got a value of 0.77, indicating acceptable reliability for measuring personal hygiene practices. After considering students feedback, we have included both three-point and four-point Likert scales response format to quantify the frequency of hygiene practice. Items on a three-point scale were scored as (i) "Always" = 2, "Sometimes" = 1, "Never" = 0; (ii) "Daily" = 2, "Weekly" = 1, "Monthly" = 0. For the four-point scale, responses were coded as "Once a week" = 2, "Once in 15 days" = 1, "Once a month" = 1, and "Once more than one month duration" = 0. The total hygiene score ranged from 0 to 36, with higher scores indicating better hygiene practices. Participants were classified into three hygiene categories: poor (0–17), moderate (18–26), and good (27–36).

**Figure 1:**
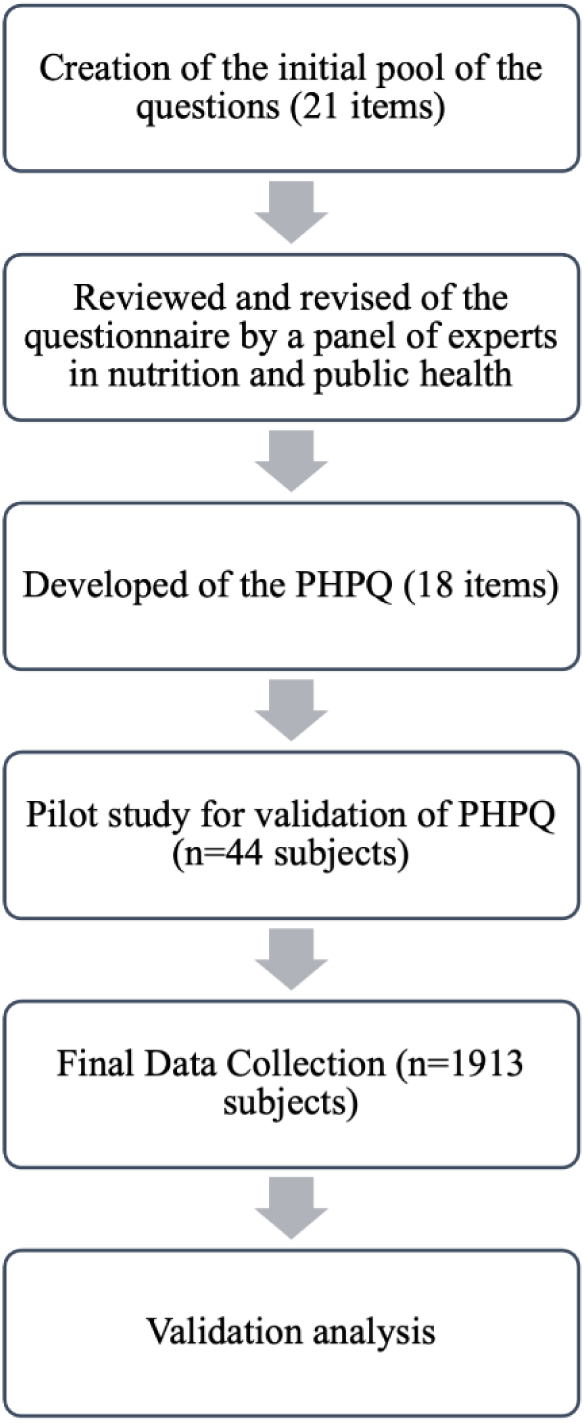
Flowchart of PHPQ development and validation process

### Statistical Analysis

We described continuous socio-demographic variables using mean, standard deviation and range and categorical variables using frequency and percentage. We used chi-square tests (chi2) to determine any statistically significant association between sociodemographic variables, depression and personal hygiene scores. An ordered logistic regression analysis was used to analyze how depression and sociodemographic factors affect personal hygiene practice. We adjusted this logistics regression analysis to account for potential confounders. Cronbach’s alpha was used to test the reliability of the CES-D scale. We used Cronbach’s alpha for reliability test and Exploratory Factor Analysis (EFA) to test the validity of our newly developed personal hygiene practice/behavior questionnaire. All statistical analyses were done using Stata 18.

## Results

### Participants’ characteristics

The total number of participants in this study was 1,913 with a mean age of 21.99 years (SD=1.61) and an age range of 18 to 33 years **(Table 1)**. Our sample included slightly more males (52.17%) than females (47.83%). The distribution of students between public (50.65%) and private universities (49.35%) was nearly equal. Among study majors, engineering students comprised the largest group (39.57%), followed by those from other disciplines (38.63%), life sciences (13.80%), and medical fields (8.0%).

**Table 1:** Socio-demographic information of the participants.

| Variables | Category | n (%) |
| --- | --- | --- |
| Age (in years) | Mean ± SD | 21.99 ± 1.61 |
|  | Range | 18-33 |
| Gender | Female | 915 (47.83) |
|  | Male | 998 (52.17) |
| Type of University | Public | 969 (50.65) |
|  | Private | 944 (49.35) |
| Study Major | Medical | 153 (8.0) |
|  | Life Sciences | 264 (13.80) |
|  | Engineering | 757 (39.57) |
|  | Others | 739 (38.63) |
| Study Level | First Year | 451 (23.58) |
|  | Second Year | 779 (40.72) |
|  | Third Year | 434 (22.69) |
|  | Fourth Year | 249 (13.02) |
| Accommodation | Home | 732 (38.26) |
|  | Provided by the University | 530 (27.71) |
|  | Privately Managed | 651 (34.03) |
| **Education of Mother** | Illiterate | 40 (2.09) |
|  | Primary | 189 (9.88) |
|  | Secondary | 712 (37.22) |
|  | Undergraduate | 536 (28.02) |
|  | Graduate and post-graduate | 436 (22.79) |
| **Education of Father** | Illiterate | 41 (2.19) |
|  | Primary | 120 (6.27) |
|  | Secondary | 413 (21.59) |
|  | Undergraduate | 527 (27.55) |
|  | Graduate and post-graduate | 812 (42.45) |
| **Family Income** | Lowest (Q1) | 526 (27.50) |
|  | Second (Q2) | 593 (31.00) |
|  | Third (Q3) | 344 (17.98) |
|  | Highest (Q4) | 450 (23.52) |

All categorical variables are expressed in percentage (%)
All continuous variables are expressed in mean and standard deviation.

Regarding study level, second-year students represented the largest proportion (40.72%), followed by first-year (23.58%), third-year (22.69%), and fourth-year students (13.02%). Place of residence varied among students, with 38.26% living at home, 27.71% residing in university-provided housing, and 34.03% in private accommodation.

We also collected data on parental education levels and found that 37.22% of students’ mothers and 21.59% of fathers had attained secondary education and 42.45% of fathers and 22.79% of mothers had graduate or postgraduate degrees. We categorized the family income of our participants into income quartiles, where 27.50% in the lowest quartile (Q1), 31.00% in the second quartile (Q2), 17.98% in the third quartile (Q3), and 23.52% in the highest quartile (Q4).

### Socio-demographic variables, depression and personal hygiene

We analyzed the association between socio-demographic factors and both depression risk (CES-D score categories) and personal hygiene practice categories among university students **(Table 2)**. Our findings show that gender significantly influences both depression risk and personal hygiene practices. A higher proportion of females (79.9%) were at risk of depression compared to males (73.9%) (χ² = 9.77, p = 0.002). Females demonstrated better personal hygiene practices, with 90.1% classified as having good hygiene compared to 75.0% of males (χ² = 74.73, p < 0.001).

**Table 2:** Association of Socio-Demographic variables with CES-D Category and Personal Hygiene Practice Category.

| Socio-Demographic Variables | CES-D Category | | | $\chi^2$ | p-value | Personal Hygiene Practice | | | | $\chi^2$ | p-value |
| --- | --- | --- | --- | --- | --- | --- | --- | --- | --- | --- | --- |
|  | Not at risk<br>(n, %) | At risk<br>(n, %) | Total<br>(n, %) |  |  | Poor | Moderate | Good | Total<br>(n, %) |  |  |
| Gender |  |  |  |  |  |  |  |  |  |  |  |
| Female | 184<br>(20.1) | 731<br>(79.9) | 915<br>(100) | 9.77 | **0.002** | 4<br>(0.4) | 87 (9.5) | 824<br>(90.1) | 915<br>(100) | 74.73 | **<0.001** |
| Male | 261<br>(26.1) | 737<br>(73.9) | 998<br>(100) |  |  | 16<br>(1.6) | 234<br>(23.4) | 748<br>(75.0) | 998<br>(100) |  |  |
| University Type |  |  |  |  |  |  |  |  |  |  |  |
| Public | 230<br>(23.7) | 739<br>(76.3) | 969<br>(100) | 0.25 | 0.619 | 7<br>(0.7) | 175<br>(18.1) | 787<br>(81.2) | 969<br>(100) | 4.10 | 0.129 |
| Private | 215<br>(22.8) | 729<br>(77.2) | 944<br>(100) |  |  | 13<br>(1.4) | 146<br>(15.5) | 785<br>(83.1) | 944<br>(100) |  |  |
| Study Major |  |  |  |  |  |  |  |  |  |  |  |
| Medical | 45<br>(29.4) | 108<br>(70.6) | 153<br>(100) | 3.78 | 0.286 | 2<br>(1.3) | 26 (17.0) | 125<br>(81.7) | 153<br>(100) | 6.83 | 0.337 |
| Life Sciences | 57<br>(21.6) | 207<br>(78.4) | 264<br>(100) |  |  | 2<br>(0.8) | 37 (14.0) | 225<br>(85.2) | 264<br>(100) |  |  |
| Engineering | 175<br>(23.1) | 582<br>(76.9) | 757<br>(100) |  |  | 5<br>(0.7) | 142<br>(18.8) | 610<br>(80.5) | 757<br>(100) |  |  |
| Others | 168<br>(22.7) | 571<br>(77.3) | 739<br>(100) |  |  | 11<br>(1.5) | 116<br>(15.7) | 612<br>(82.8) | 739<br>(100) |  |  |
| Study Level |  |  |  |  |  |  |  |  |  |  |  |
| First Year | 108<br>(23.9) | 343<br>(76.1) | 451<br>(100) | 1.36 | 0.714 | 3<br>(0.7) | 58 (12.9) | 390<br>(86.5) | 451<br>(100) | 12.43 | 0.053 |
| Second Year | 171<br>(22.0) | 608<br>(78.0) | 779<br>(100) |  |  | 13<br>(1.7) | 135<br>(17.3) | 631<br>(81.0) | 779<br>(100) |  |  |
| Third Year | 104<br>(24.0) | 330<br>(76.0) | 434<br>(100) |  |  | 3<br>(0.7) | 79 (18.2) | 352<br>(81.1) | 434<br>(100) |  |  |
| Fourth Year | 62<br>(24.9) | 187<br>(75.1) | 249<br>(100) |  |  | 1<br>(0.4) | 49 (19.7) | 199<br>(79.9) | 249<br>(100) |  |  |
| Accommodation |  |  |  |  |  |  |  |  |  |  |  |
| Home | 110<br>(20.8) | 420<br>(79.2) | 530<br>(100) | 8.26 | 0.016 | 5<br>(0.9) | 115<br>(21.7) | 410<br>(77.4) | 530<br>(100) | 36.82 | <0.001 |
| Provided by the University | 139<br>(21.3) | 512<br>(78.7) | 651<br>(100) |  |  | 7<br>(1.1) | 131<br>(20.1) | 513<br>(78.8) | 651<br>(100) |  |  |
| Privately Managed | 196<br>(26.8) | 536<br>(73.2) | 732<br>(100) |  |  | 8<br>(1.1) | 75 (10.2) | 649<br>(88.6) | 732<br>(100) |  |  |
| Education of Mother |  |  |  |  |  |  |  |  |  |  |  |
| Illiterate | 8<br>(20.0) | 32<br>(80.0) | 40<br>(100) | 3.34 | 0.503 | 0<br>(0.0) | 14 (35.0) | 26<br>(65.0) | 40<br>(100) | 22.48 | 0.004 |
| Primary | 37<br>(19.6) | 152<br>(80.4) | 189<br>(100) |  |  | 2<br>(1.1) | 46 (24.3) | 141<br>(74.6) | 189<br>(100) |  |  |
| Secondary | 177<br>(24.9) | 535<br>(75.1) | 712<br>(100) |  |  | 8<br>(1.1) | 112<br>(15.7) | 592<br>(83.2) | 712<br>(100) |  |  |
| Undergraduate | 128<br>(23.9) | 408<br>(76.1) | 536<br>(100) |  |  | 3<br>(0.6) | 86 (16.0) | 447<br>(83.4) | 536<br>(100) |  |  |
| Graduate and post-graduate | 95<br>(21.8) | 341<br>(78.2) | 436<br>(100) |  |  | 7<br>(1.6) | 63 (14.4) | 366<br>(84.0) | 436<br>(100) |  |  |
| Education of Father |  |  |  |  |  |  |  |  |  |  |  |
| Illiterate | 9<br>(21.9) | 32<br>(78.1) | 41<br>(100) | 0.27 | 0.991 | 0<br>(0.0) | 11 (26.8) | 30<br>(73.2) | 41<br>(100) | 16.35 | 0.038 |
| Primary | 29<br>(24.2) | 91<br>(75.8) | 120<br>(100) |  |  | 1<br>(0.8) | 33 (27.5) | 86<br>(71.7) | 120<br>(100) |  |  |
| Secondary | 97<br>(23.5) | 316<br>(76.5) | 413<br>(100) |  |  | 4<br>(1.0) | 65 (15.7) | 344<br>(83.3) | 413<br>(100) |  |  |
| Undergraduate | 119<br>(22.6) | 408<br>(77.4) | 527<br>(100) |  |  | 5<br>(0.9) | 75 (14.2) | 447<br>(84.8) | 527<br>(100) |  |  |
| Graduate and<br>post-graduate | 191<br>(23.5) | 621<br>(76.5) | 812<br>(100) |  |  | 10<br>(1.2) | 137<br>(16.9) | 665<br>(81.9) | 812<br>(100) |  |  |
| Family Income |  |  |  |  |  |  |  |  |  |  |  |
| Lowest (Q1) | 116<br>(22.1) | 410<br>(77.9) | 526<br>(100) | 3.93 | 0.270 | 5<br>(0.9) | 112<br>(21.3) | 409<br>(77.8) | 526<br>(100) | 13.13 | 0.040 |
| Second (Q2) | 133<br>(22.4) | 460<br>(77.6) | 593<br>(100) |  |  | 5<br>(0.8) | 91 (15.4) | 497<br>(83.8) | 593<br>(100) |  |  |
| Third (Q3) | 94<br>(27.3) | 250<br>(72.7) | 344<br>(100) |  |  | 3<br>(0.9) | 46 (13.4) | 295<br>(85.8) | 344<br>(100) |  |  |
| Highest (Q4) | 102<br>(22.7) | 348<br>(77.3) | 450<br>(100) |  |  | 7<br>(1.6) | 72 (16.0) | 371<br>(82.4) | 450<br>(100) |  |  |

Accommodation type was significantly associated with depression and personal hygiene. Students living in privately managed accommodations had a lower risk of depression (73.2%) compared to those living at home (79.2%) or in university-provided housing (78.7%) (χ² = 8.26, p = 0.016). They also showed better hygiene practices, with 88.6% categorized as having good hygiene, compared to 77.4% and 78.8% for home and university-provided housing, respectively (χ² = 36.82, p < 0.001).

Parental education level also influenced personal hygiene, students whose mothers had lower education levels exhibiting poorer hygiene (χ² = 22.48, p = 0.004). Similarly, father’s education showed a significant association with hygiene practices (χ² = 16.35, p = 0.038), though it did not significantly impact depression risk (p = 0.991).

Family income was also significantly related to personal hygiene practices (χ² = 13.13, p = 0.04). Students from the lowest income quartile (Q1) had poorer hygiene practices than those from higher income groups, though income did not show a significant relationship with depression risk (p = 0.27).

Other socio-demographic variables, such as university type, subject major, and level of study did not show statistically significant associations with depression risk or personal hygiene practices.

We conducted another chi-square test to identify any association between depression (not at risk and at risk) and personal hygiene practice (poor, moderate and good). From the **Table-3**, we can see a statistically significant association between depression and personal hygiene practice ( χ² = 31.45, p < 0.001). The majority (91.0%) of the students who were not at risk of depression had good hygiene practices, 8.8% had moderate and 0nly 0.2% had poor hygiene practices. On the other hand, 79.5% of students who were at risk of depression had good hygiene practices, 19.2% had moderate and 1.3% had poor hygiene practices. The chi-square results show that students at risk of depression were less likely to maintain good hygiene practices and more likely to fall into moderate or poor hygiene categories compared to those who were not at risk of depression.

**Table 3:** Association between Depression and Personal Hygiene Practice among University Students.

| CES-D<br>Score<br>Category | Poor<br>(n,%) | Moderate (n,%) | Good (n,%) | Total (n,%) | $\chi^2$ | df | p-value |
| --- | --- | --- | --- | --- | --- | --- | --- |
| Not at risk | 1 (0.2) | 39 (8.8) | 405 (91.0) | 445 (100) | 31.45 | 2 | <0.001 |
| At risk | 19 (1.3) | 282 (19.2) | 1,167 (79.5) | 1,468 (100) |  |  |  |
| Total | 20 (1.0) | 321 (16.8) | 1,572 (82.2) | 1,913 (100) |  |  |  |

We conducted an ordered logistic regression analysis to explore the impact of depression and socio-demographic factors on personal hygiene practices among university students **(Table 4)**. Students at risk of depression had 65% lower odds of better hygiene practices compared to those not at risk (OR = 0.36, p < 0.001). Male students were 68% less likely to have higher hygiene scores than females (OR = 0.32, p < 0.001). Accommodation type also showed a significant association with personal hygiene practices. Students living in privately managed accommodations had nearly 2 times higher odds of better hygiene practices compared to those living at home (OR = 1.99, 95% CI: 1.42–2.80, p < 0.001). Second-year students were 34% less likely to practice better hygiene compared to first-year students (OR = 0.66, 95% CI: 0.47–0.94, p = 0.022). However, no significant differences were observed for third- and fourth-year students.

**Table 4:**
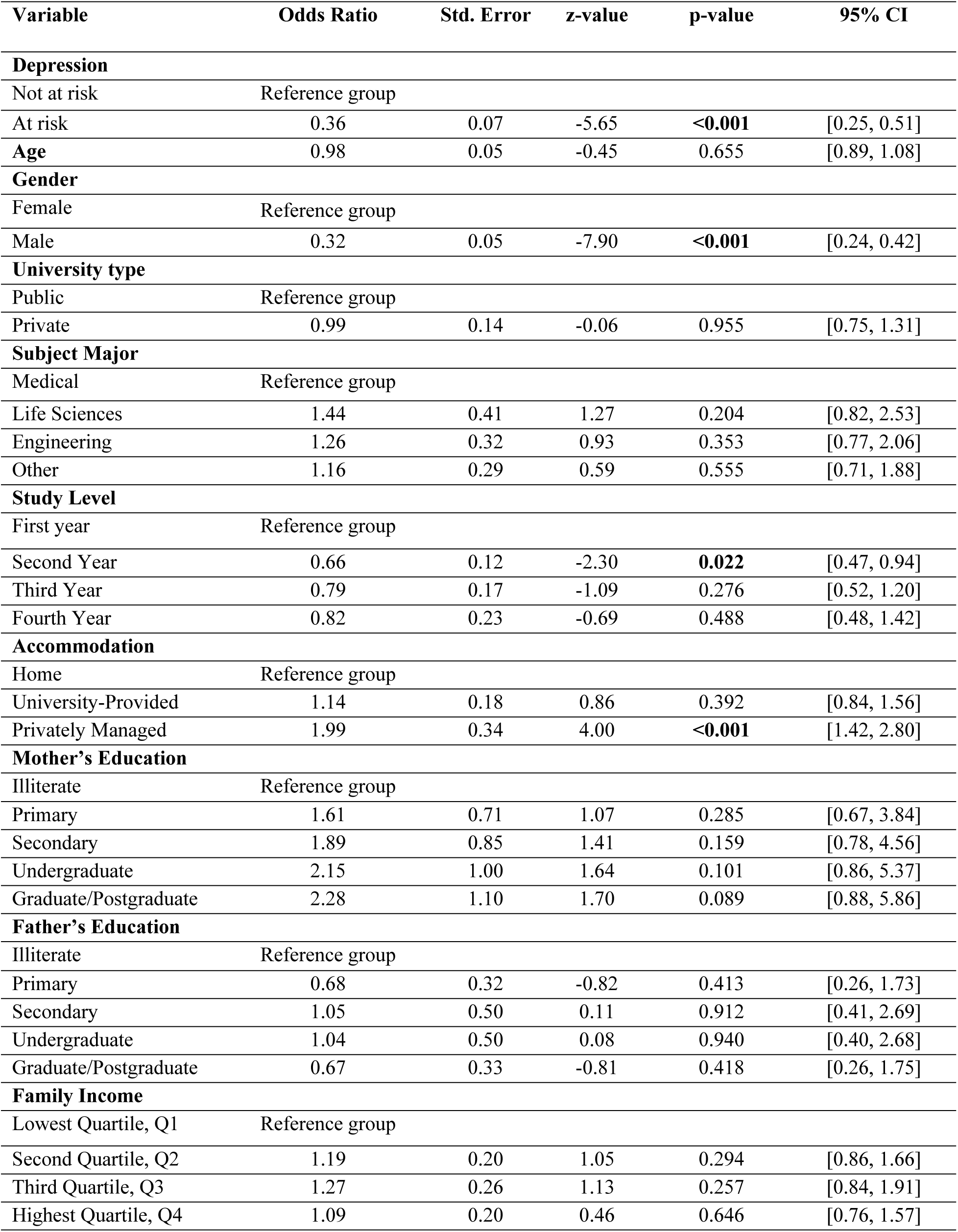
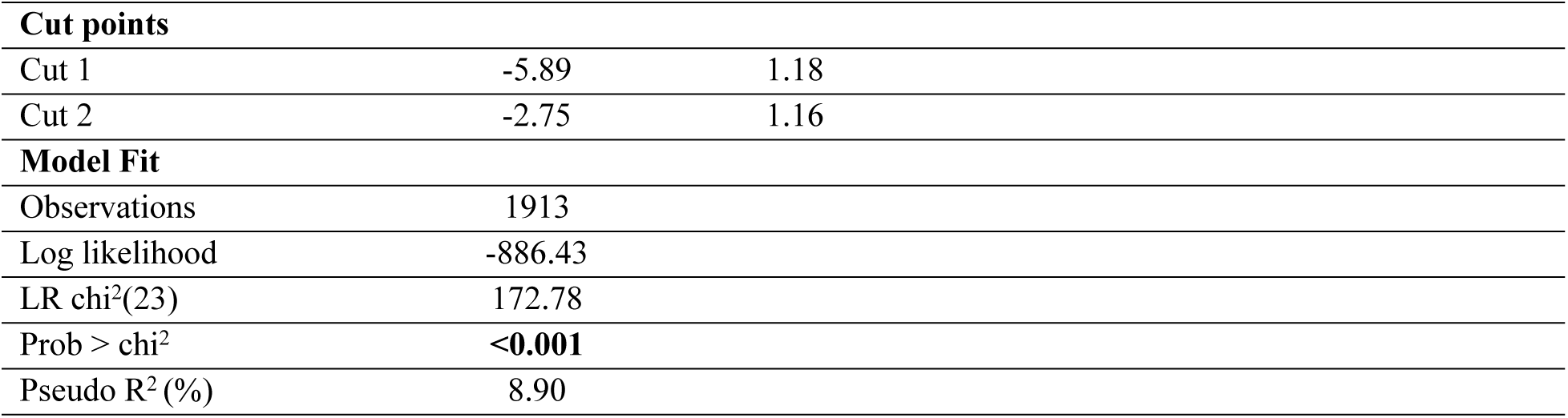
Ordered Logistic Regression Analysis of Variables Influencing Personal Hygiene Practice.

| Variable | Odds Ratio | Std. Error | z-value | p-value | 95% CI |
| --- | --- | --- | --- | --- | --- |
| **Depression** |  |  |  |  |  |
| Not at risk | Reference group |  |  |  |  |
| At risk | 0.36 | 0.07 | -5.65 | <0.001 | [0.25, 0.51] |
| **Age** | 0.98 | 0.05 | -0.45 | 0.655 | [0.89, 1.08] |
| **Gender** |  |  |  |  |  |
| Female | Reference group |  |  |  |  |
| Male | 0.32 | 0.05 | -7.90 | <0.001 | [0.24, 0.42] |
| **University type** |  |  |  |  |  |
| Public | Reference group |  |  |  |  |
| Private | 0.99 | 0.14 | -0.06 | 0.955 | [0.75, 1.31] |
| **Subject Major** |  |  |  |  |  |
| Medical | Reference group |  |  |  |  |
| Life Sciences | 1.44 | 0.41 | 1.27 | 0.204 | [0.82, 2.53] |
| Engineering | 1.26 | 0.32 | 0.93 | 0.353 | [0.77, 2.06] |
| Other | 1.16 | 0.29 | 0.59 | 0.555 | [0.71, 1.88] |
| **Study Level** |  |  |  |  |  |
| First year | Reference group |  |  |  |  |
| Second Year | 0.66 | 0.12 | -2.30 | 0.022 | [0.47, 0.94] |
| Third Year | 0.79 | 0.17 | -1.09 | 0.276 | [0.52, 1.20] |
| Fourth Year | 0.82 | 0.23 | -0.69 | 0.488 | [0.48, 1.42] |
| **Accommodation** |  |  |  |  |  |
| Home | Reference group |  |  |  |  |
| University-Provided | 1.14 | 0.18 | 0.86 | 0.392 | [0.84, 1.56] |
| Privately Managed | 1.99 | 0.34 | 4.00 | <0.001 | [1.42, 2.80] |
| **Mother's Education** |  |  |  |  |  |
| Illiterate | Reference group |  |  |  |  |
| Primary | 1.61 | 0.71 | 1.07 | 0.285 | [0.67, 3.84] |
| Secondary | 1.89 | 0.85 | 1.41 | 0.159 | [0.78, 4.56] |
| Undergraduate | 2.15 | 1.00 | 1.64 | 0.101 | [0.86, 5.37] |
| Graduate/Postgraduate | 2.28 | 1.10 | 1.70 | 0.089 | [0.88, 5.86] |
| **Father's Education** |  |  |  |  |  |
| Illiterate | Reference group |  |  |  |  |
| Primary | 0.68 | 0.32 | -0.82 | 0.413 | [0.26, 1.73] |
| Secondary | 1.05 | 0.50 | 0.11 | 0.912 | [0.41, 2.69] |
| Undergraduate | 1.04 | 0.50 | 0.08 | 0.940 | [0.40, 2.68] |
| Graduate/Postgraduate | 0.67 | 0.33 | -0.81 | 0.418 | [0.26, 1.75] |
| **Family Income** |  |  |  |  |  |
| Lowest Quartile, Q1 | Reference group |  |  |  |  |
| Second Quartile, Q2 | 1.19 | 0.20 | 1.05 | 0.294 | [0.86, 1.66] |
| Third Quartile, Q3 | 1.27 | 0.26 | 1.13 | 0.257 | [0.84, 1.91] |
| Highest Quartile, Q4 | 1.09 | 0.20 | 0.46 | 0.646 | [0.76, 1.57] |

|  |  |  |
| --- | --- | --- |
| **Cut points** |  |  |
| Cut 1 | -5.89 | 1.18 |
| Cut 2 | -2.75 | 1.16 |
| **Model Fit** |  |  |
| Observations | 1913 |  |
| Log likelihood | -886.43 |  |
| LR chi<sup>2</sup>(23) | 172.78 |  |
| Prob > chi<sup>2</sup> | **<0.001** |  |
| Pseudo R<sup>2</sup> (%) | 8.90 |  |

Although higher maternal education showed a positive association with better hygiene practices, the results were not statistically significant. For example, students whose mothers had a postgraduate education had 2.28 times higher odds of better hygiene practices than students whose mothers were illiterate, but the effect did not reach significance (OR = 2.28, 95% CI: 0.88–5.86, p = 0.089). Paternal education, university type, subject major, age, and family income did not show statistically significant associations with personal hygiene practices, as their 95% confidence intervals included 1.

By adjusting for potential confounders such as age, gender, income, education, and accommodation, we strengthened the model’s reliability. The model was statistically significant (χ²= 172.78, p < 0.001), and explained approximately 8.9% of the variance in personal hygiene practices (Pseudo R² = 0.0888). The small effect sizes suggest that the additional factors (e.g. awareness on hygiene related diseases, individual belief about cleanliness, availability of hygiene facilities, social and cultural influences etc.) beyond those included in this analysis may contribute to personal hygiene behavior.

### Reliability and validity of the personal hygiene practice/behavior questionnaire

A Cronbach’s alpha over 0.70 indicates that the data are reliable and consistently measure a construct (15). In our study the overall Cronbach’s alpha for the questionnaire we used to assess personal hygiene practice was 0.83 which means good internal consistency **(Table 5)**. For most of our items, the item-test correlation ranging from 0.23 to 0.63 exceeded 0.40 which indicated that there was a strong association between each item and the overall scale. All item-rest correlations (ranging from 0.16 to 0.56) except the “cutting nails” item had contributions to the personal hygiene practice scale’s internal consistency. The average inter-item covariance of 0.06 demonstrated consistency across the 18-item of our questionnaire.

**Table 5:** Internal Reliability of Personal Hygiene Practice Questionnaire.

| Items | Item-test correlation | Item-rest Correlation | Average interitem covariance | alpha ($\alpha$) |
| --- | --- | --- | --- | --- |
| Handwash before eating | 0.45 | 0.39 | 0.06 | 0.83 |
| Handwash after eating | 0.48 | 0.42 | 0.06 | 0.83 |
| Handwash with soap after toilet use | 0.61 | 0.52 | 0.06 | 0.82 |
| Handwash after blowing nose | 0.53 | 0.44 | 0.06 | 0.83 |
| Handwash after touching animal | 0.61 | 0.51 | 0.05 | 0.82 |
| Handwash after touching private parts | 0.63 | 0.56 | 0.06 | 0.82 |
| Brushing teeth | 0.48 | 0.43 | 0.06 | 0.83 |
| Bathing | 0.51 | 0.45 | 0.06 | 0.83 |
| Wear clean clothes | 0.58 | 0.51 | 0.06 | 0.82 |
| Change underwear | 0.61 | 0.55 | 0.06 | 0.82 |
| Removing unwanted hair | 0.53 | 0.45 | 0.06 | 0.83 |
| Wash hair | 0.52 | 0.44 | 0.06 | 0.83 |
| Handkerchief/tissue use after blowing nose | 0.62 | 0.54 | 0.06 | 0.82 |
| Cutting nail | 0.23 | 0.16 | 0.06 | 0.84 |
| Changing bedsheet | 0.49 | 0.39 | 0.06 | 0.83 |
| Changing pillow cover | 0.48 | 0.37 | 0.06 | 0.83 |
| Dusting own room | 0.47 | 0.36 | 0.06 | 0.83 |
| Mopping own room | 0.47 | 0.36 | 0.06 | 0.83 |
| Total Scale |  |  | 0.06 | 0.83 |

**Figure 2:**
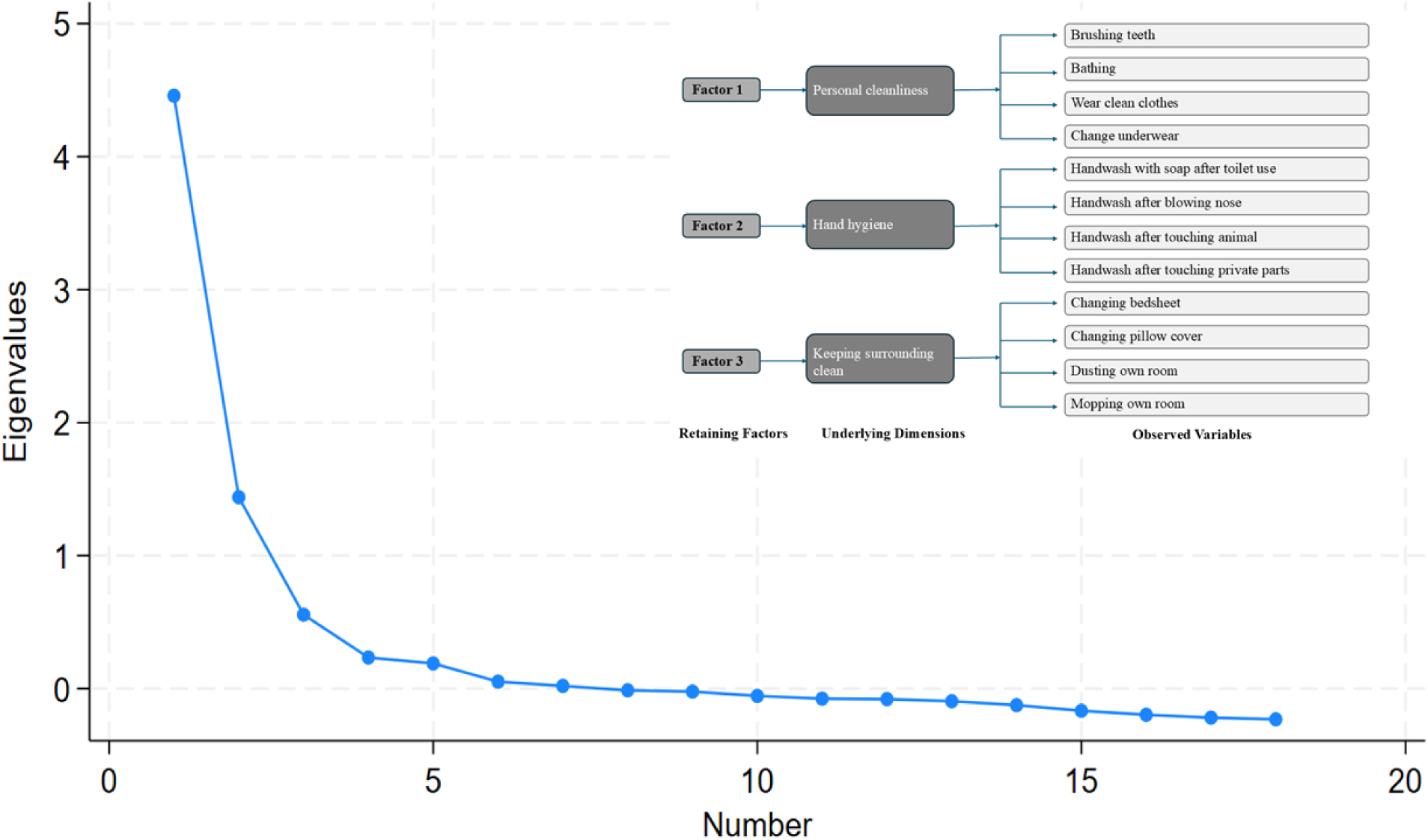
Factor analysis by scree plot

We did Exploratory Factor Analysis (EFA) to test the validity of the personal hygiene questionnaire. The results of the EFA supported the construct validity of the 18-item personal hygiene practice questionnaire **(Table 6)**. We got overall 0.89 value from the Kaiser– Meyer– Olkin (KMO) test and χ²(153) = 8924.81, p < 0.001 from the Bartlett’s test of sphericity. The analysis retained three factors based on eigenvalues (>1), the scree plot and proportion of explained variance. Factor 1 is comprised of four items that are related to personal cleanliness practices (brushing teeth, bathing, wearing clean clothes and changing underwear), Factor 2 was associated with four items that were related to hand hygiene practice dimension, and Factor 3 was composed of another four items that reflected a dimension focused on keeping the surrounding clean. From table 7, we found that factor 1 had a factor loading ranging from 0.59 to 0.64 with a Cronbach’s alpha of 0.75 and relatively a low uniqueness value (0.56 to 0.58), factor 2 had loadings from 0.45 to 0.60, an alpha of 0.72 and uniqueness value ranging from 0.53 to 0.69, and factor 3, reflecting surrounding cleanliness with loadings between 0.50 and 0.67, an alpha of 0.72 and uniqueness value of 0.53 to 0.72. Overall, a factor loading ≥ 0.45 tells us that each item is well associated with the respective factors. Most of the item’s uniqueness values fell below 0.70 in the EFA analysis which indicated that the extracted factors explained a considerable portion of variance in the personal hygiene questionnaire items. From these findings, we could say that the questionnaire was a valid and reliable tool for assessing personal hygiene practice among university students.

**Table 6:**
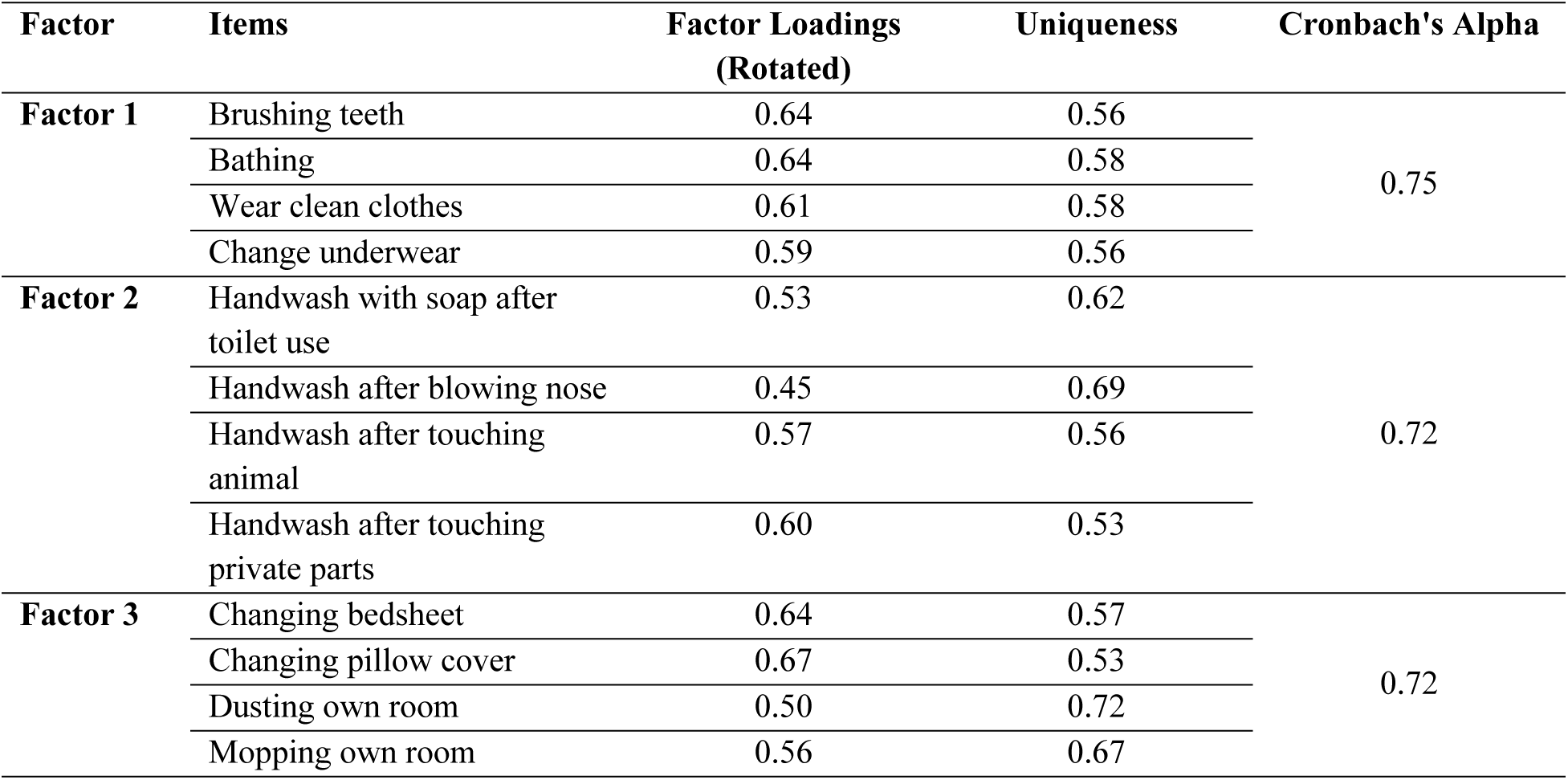
Exploratory Factor Analysis of the Personal Hygiene Practice Questionnaire.

| Factor | Items | Factor Loadings<br>(Rotated) | Uniqueness | Cronbach's Alpha |
| --- | --- | --- | --- | --- |
| **Factor 1** | Brushing teeth | 0.64 | 0.56 | 0.75 |
|  | Bathing | 0.64 | 0.58 |  |
|  | Wear clean clothes | 0.61 | 0.58 |  |
|  | Change underwear | 0.59 | 0.56 |  |
| **Factor 2** | Handwash with soap after toilet use | 0.53 | 0.62 | 0.72 |
|  | Handwash after blowing nose | 0.45 | 0.69 |  |
|  | Handwash after touching animal | 0.57 | 0.56 |  |
|  | Handwash after touching private parts | 0.60 | 0.53 |  |
| **Factor 3** | Changing bedsheet | 0.64 | 0.57 | 0.72 |
|  | Changing pillow cover | 0.67 | 0.53 |  |
|  | Dusting own room | 0.50 | 0.72 |  |
|  | Mopping own room | 0.56 | 0.67 |  |

## Discussion

The aim of our study was to assess the relationship between depression and personal hygiene practices among university students along with the validation of our newly developed personal hygiene questionnaire in order to make this self-rated questionnaire. In our study, we identified a statistically significant association between depression and personal hygiene practice. We also found a high prevalence of depression risk among male and female students but comparing with male, female students had a higher-level depression risk. In terms of personal hygiene, female students maintain good personal hygiene practice compared with male. Along with gender, depression risk among students is associated with accommodation of students whereas personal hygiene practices are associated with level of study, accommodation, parental education and family income status.

The findings of our study about the prevalence of depression risk among university students are higher than most recently published studies among students of Bangladesh. Though these studies used different scale of depression including the WHO-5 Well-Being Index (WHO-5) (16), and 9-item Patient Health Questionnaire (PHQ-9) (10,17), and Depression, Anxiety, and Stress Scale (DASS-42) scale (18); the authors from these studies stated the prevalence of depression among university students ranging from 42% to 52%. These studies revealed a higher prevalence of depression level among female students compared with male (10,16–18). However, another study among first year university students reported a higher prevalence of depressive symptoms among male (50.4%) compared with female (49.6%)(19) inconsistent with the present study. Another study using DASS-21 scale among public university students reported a non-significant higher prevalence of depression level among students who stayed at hall/mess (20). The differences of depression prevalence regarding accommodation status in Hossain, Alam and Masum’s study and our study could be due to inclusion of only public university students whereas we included both public and private university students.

Depression is another important factor influencing personal hygiene practices explored by the present study. In Bangladesh, a limited number of studies evaluated personal hygiene practices among university students. Beyond the school-based WASH related articles, some studies evaluated the hand washing practices at university settings along with their knowledge and attitudes regarding this (21,22). And to our best knowledge, no quantitative study was found to evaluate the personal hygiene practices considering other personal hygiene related variables as well as the impact of mental health on personal hygiene practices. This shortage of literature may limit the scope of comparing the findings of our study with others. The prevalence of practicing good personal hygiene was higher among female students than male in our study. Similar with present study, female students tended to greater use soap for hand washing within their college compared with men (5,23). The study noted that personal hygiene was significantly associated with socio-economic status. Consistent with present study, students belonging from middle or upper-middle income family had good personal hygiene practices as they have greater access to hygiene information (i.e., social media, newspaper, and other media exposure) (24,25). Living place is considered as another reason for good personal hygiene among university students. Due to high economic growth over the last two decades in Bangladesh, sanitation and hygiene facilities among school and university settings are increased rapidly. The demands of hygiene practices are also increased. To cover up the demands, the government of Bangladesh announced a program named “Sanitation for All by 2010” and all educational institutions are trying to comply with the national goals (26). In our study, the percentage of maintaining good personal hygiene among university is quite high in both male and female, which may be due to practices of personal hygiene during the COVID-19 period.

The present study revealed that the risk of depression among university students may reduce good personal hygiene practices. A recent study among health professionals found that depressive persons had low level of standards regarding personal hygiene and grooming (27) consistent with the present study. However, low-economic status, lack of social activity and support and lack of proper vocational and academic opportunities have been linked with depression among individuals (28). A recent scoping review revealed that depressed individuals were less likely to wash their hands. They did not have any guilt or not have any intension to wash their hands with soap (29). According to the definition of depression, individual’s daily life activity at work or schools are impaired by depression (30). However, another study conducted among children found inverse relationship between hand washing and depression (31). Students reporting academic pressure emerged another predictive factor of depression which may lead to lower personal hygiene practices. Education related to personal hygiene may also influence good personal hygiene practices. We have found that engineering students have better personal hygiene practices compared with other educational background students. In opposition to our findings, students from health and life sciences background had better knowledge, attitude and practices regarding personal hygiene compared with engineering or other background students (32).

As we previously mentioned, literatures about personal hygiene practices among university is limited and no study yet carried out at Bangladesh settings, we had to develop and validate a new questionnaire tool to reduce the gap in literature for assessing the personal hygiene practices among students. EPA analysis suggested that this questionnaire has a correlated three factor structures. From the measurement of internal consistency and construct validity, this questionnaire has been demonstrated as a reliable measure for personal hygiene practices among university students.

Nevertheless, this study has been limited by several factors. Firstly, the pseudo-R-squared value for the regression model is much lower (0.089) indicating that the model can explain only a small portion of variance in the dependent variables although the findings are significant. Others unmeasured variables might influence our findings such as social welfare, personal care facility, social support, availability and accessibility to hygiene facilities, self-esteem, religious and cultural beliefs and practices and motivation. Secondly, data were collected from universities in Dhaka, a central hub for higher education in Bangladesh, which may limit the generalizability to the broader university student population nationwide. Thirdly, the temporal stability of this questionnaire was not assessed. And the weight of some items in loadings 2 and 3 were considered low or moderate. And lastly, we have used web-based data collection methods which may limit the number of participations as it can access to those who have internet. This may also introduce bias in sampling.

Despite these limitations, it is worth mentioning some strengths of our study. First, this is the first study in Bangladesh which evaluates the effect of depression on personal hygiene among university students. Secondly, we have used a newly developed and validate personal hygiene practices questionnaire with good reliability and validity score which can be used in future (after additional item inclusion) for assessing personal hygiene at university settings. Thirdly, we have included a diverse range of participations from different disciplines, study year, semester, residence to maximize the variation.

## Conclusion

This study provides a comprehensive analysis of the relationship between depression risk and personal hygiene practices among university students, revealing critical insights into the mental and physical health of this demographic. Gender and accommodation type were significant determinants; females displayed superior personal cleanliness habits, whilst students living in privately managed lodgings showed a reduced incidence of depression and higher hygiene standards. These findings emphasize the necessity for educational institutions to focus mental health programs and hygiene instruction. It is imperative to implement regular mental health screenings to identify at-risk students early and provide appropriate interventions. Universities should also develop workshops focused on personal hygiene education, emphasizing its connection to mental well-being.

Future initiatives may involve establishing peer support networks to cultivate a sense of community, so alleviating feelings of isolation that can intensify depression. Involving students in initiatives that advocate for hygiene awareness and mental health services can contribute to the destigmatization of these concerns. Moreover, subsequent research should investigate the socio-economic determinants affecting personal cleanliness and mental health, as this study predominantly concentrated on demographic characteristics. Exploring the impact of cultural beliefs and access to hygiene facilities will provide a more nuanced understanding of these relationships. Addressing the intertwined issues of mental health and personal hygiene through targeted interventions can lead to improved health outcomes. By fostering a supportive and informed university environment, we can enhance the overall well-being of students, equipping them with the tools necessary for both academic success and personal health.

## Data Availability

All relevant data are within the manuscript. Dataset generated for this manuscript will be made available upon request from corresponding author.

## Acknowledgement

We are grateful to all the participants and appreciate the support of all enumerators. We are also thankful to all the faculty members who provided their insights into developing the questionnaire to assess personal hygiene practice.

## References

1. Islam MA, Barna SD, Raihan H, Khan MNA, Hossain MT. Depression and anxiety among university students during the COVID-19 pandemic in Bangladesh: A web-based cross-sectional survey. PLoS One. 2020;15(8):e0238162.

2. Islam S, Akter R, Sikder T, Griffiths MD. Prevalence and factors associated with depression and anxiety among first-year university students in Bangladesh: a cross-sectional study. Int J Ment Health Addict. 2020;1–14.

3. Beiter R, Nash R, McCrady M, Rhoades D, Linscomb M, Clarahan M, et al. The prevalence and correlates of depression, anxiety, and stress in a sample of college students. J Affect Disord. 2015;173:90–6.

4. Alim SMAHM, Rabbani MG, Karim E, Mullick MSI, Al Mamun A, Khan MZR. Assessment of depression, anxiety and stress among first year MBBS students of a public medical college, Bangladesh. Bangladesh Journal of Psychiatry. 2015;29(1):23–9.

5. Al-Rifaai JM, Al Haddad AM, Qasem JA. Personal hygiene among college students in Kuwait: A Health promotion perspective. J Educ Health Promot. 2018;7(1):92.

6. Twumwaa H, Asumang B, Imoro ZA, Kpordze SW. Toothbrush and towel handling and their microbial quality: the case of students of university for development studies, nyankpala campus, GHANA. Afr J Infect Dis. 2021;15(1):41–6.

7. Schulz R, Sherwood PR. Physical and mental health effects of family caregiving. J Soc Work Educ. 2008;44(sup3):105–13.

8. Vieira F da ST, Muraro AP, Rodrigues PRM, Sichieri R, Pereira RA, Ferreira MG. Lifestyle-related behaviors and depressive symptoms in college students. Cad Saude Publica. 2021;37:e00202920.

9. Sayeed A, Rahman MH, Hassan MN, deSteiguer A, Kundu S, Meem AE, et al. Prevalence and associated factors of depression among Bangladeshi university students: A cross-sectional study. Journal of American college health. 2023;71(5):1381–6.

10. Koly KN, Sultana S, Iqbal A, Dunn JA, Ryan G, Chowdhury AB. Prevalence of depression and its correlates among public university students in Bangladesh. J Affect Disord. 2021;282:689–94.

11. Lai FTT, Chan VKY, Li TW, Li X, Hobfoll SE, Lee TMC, et al. Disrupted daily routines mediate the socioeconomic gradient of depression amid public health crises: a repeated cross-sectional study. Australian & New Zealand Journal of Psychiatry. 2022;56(10):1320–31.

12. Sharma MK, Adhikari R, Khanal SP, Acharya D, van Teijlingen E. Do school Water, Sanitation, and Hygiene facilities affect students’ health status, attendance, and educational achievements? A qualitative study in Nepal. Health Sci Rep. 2024;7(8):e2293.

13. Jasim MM, Siddiqui K. A university in every 5.38 square kilometres. 2019 Nov 2;

14. Carleton RN, Thibodeau MA, Teale MJN, Welch PG, Abrams MP, Robinson T, et al. The center for epidemiologic studies depression scale: a review with a theoretical and empirical examination of item content and factor structure. PLoS One. 2013;8(3):e58067.

15. Sharma B. A focus on reliability in developmental research through Cronbach’s Alpha among medical, dental and paramedical professionals. Asian Pacific Journal of Health Sciences. 2016;3(4):271–8.

16. Karim R, Khan SI, Bari R, Akash MA, Chowdhury AK, Seum MAA, et al. Factors Affecting Stress and Depression in Bangladeshi Students: A Cross-sectional Study. Asian Journal of Social Health and Behavior. 2024;7(2):84–93.

17. Kundu S, Bakchi J, Al Banna MH, Sayeed A, Hasan MT, Abid MT, et al. Depressive symptoms associated with loneliness and physical activities among graduate university students in Bangladesh: findings from a cross-sectional pilot study. Heliyon. 2021;7(3).

18. Kamruzzaman M, Hossain A, Islam MA, Ahmed MS, Kabir E, Khan MN. Exploring the prevalence of depression, anxiety, and stress among university students in Bangladesh and their determinants. Clin Epidemiol Glob Health. 2024;28:101677.

19. Islam S, Akter R, Sikder T, Griffiths MD. Prevalence and factors associated with depression and anxiety among first-year university students in Bangladesh: a cross-sectional study. Int J Ment Health Addict. 2020;1–14.

20. Hossain MM, Alam MA, Masum MH. Prevalence of anxiety, depression, and stress among students of Jahangirnagar University in Bangladesh. Health Sci Rep. 2022;5(2):e559.

21. Sultana M, Mahumud RA, Sarker AR, Hossain SM. Hand hygiene knowledge and practice among university students: evidence from private universities of Bangladesh. Risk Manag Healthc Policy. 2016;13–20.

22. Hossain MS, Islam A, Fahad N, Das P, Rahaman A. Basic hygiene and sanitation practices among Bangladeshi university students. International Journal of Natural and Social Sciences. 2020;7(2):40–50.

23. Snow M, White Jr GL, Alder SC, Stanford JB. Mentor’s hand hygiene practices influence student’s hand hygiene rates. Am J Infect Control. 2006;34(1):18–24.

24. Kabir A, Roy S, Begum K, Kabir AH, Miah MS. Factors influencing sanitation and hygiene practices among students in a public university in Bangladesh. PLoS One. 2021;16(9):e0257663.

25. Rabbi SE, Dey NC. Exploring the gap between hand washing knowledge and practices in Bangladesh: a cross-sectional comparative study. BMC Public Health. 2013;13:1–7.

26. Hanchett S, Khan MH, Krieger L, Kullmann C. Sustainability of sanitation in rural Bangladesh. 2011;

27. Stewart V, Judd C, Wheeler AJ. Practitioners’ experiences of deteriorating personal hygiene standards in people living with depression in Australia: A qualitative study. Health Soc Care Community. 2022;30(4):1589–98.

28. Bagana E. Depression and social vulnerability to depression. Procedia-Social and Behavioral Sciences. 2013;78:456–60.

29. Ghassemi EY, Thorseth AH, Le Roch K, Heath T, White S. Mapping the association between mental health and people’s perceived and actual ability to practice hygiene-related behaviours in humanitarian and pandemic crises: A scoping review. PLoS One. 2023;18(12):e0286494.

30. World Health Organization (WHO). http://www.who.int/mental_health/management/depression/en/. Mental health. Management. Depression.

31. Slekiene J, Mosler HJ. Does depression moderate handwashing in children? BMC Public Health. 2018;18:1–9.

32. Barcenilla-Guitard M, Espart A. Influence of gender, age and field of study on hand hygiene in young adults: a cross-sectional study in the COVID-19 pandemic context. Int J Environ Res Public Health. 2021;18(24):13016.

